# Changed epidemiology of influenza and RSV hospitalizations after the emergence of SARS-CoV-2 in Norway, 2017 - 2024

**DOI:** 10.1101/2025.02.17.25322411

**Authors:** Håkon Bøås, Elina Seppälä, Lamprini Veneti, Jeanette Stålcrantz, Jacob Dag Berild, Jesper Dahl, Trine Hessevik Paulsen

## Abstract

**Background:** Many countries reported missing and atypical influenza and RSV seasons during the COVID-19 pandemic. Here we describe the incidence and seasonality of COVID-19, influenza, and RSV hospitalizations in Norway between 2017-2024, and the disease burden between 2022-2024.

**Methods:** Using nationwide data on ICD-10 discharge codes, procedure codes and laboratory results, we calculate the incidence of COVID-19, influenza, and RSV hospitalizations, by age group, week and season between January 2017 to April 2024, and report proportions receiving intensive care, proportion of deaths and length of stay between 2022-2024.

**Results:** The transmission of influenza and RSV was interrupted the first year of the COVID-19 pandemic and reemerged with epidemics outside of the normal seasonality in 2021/2022, after COVID-19 restrictions were removed. Between 2022-2024, COVID-19 was a greater contributor to hospitalizations than influenza and RSV, with higher mortality rate within two weeks of admission. The use of ventilatory support/intensive care admission was highest among patients hospitalized with RSV.

**Conclusion:** The transmission of influenza and RSV was interrupted during the first year of the COVID-19 pandemic, followed by an unusual seasonality. Although many hospitalizations are caused by RSV and influenza, COVID-19 was the largest contributor of these three to hospital burden in the first years with co-circulation.

## Introduction

Since the emergence of SARS-CoV-2, COVID-19 has constituted a large burden on hospitals [1], particularly among older adults and those with underlying medical conditions [2,3]. The use of non-pharmaceutical interventions (NPIs) to control SARS-CoV-2 spread also impacted the transmission and seasonality of other viral infections such as respiratory syncytial virus (RSV) and influenza virus, leading to a notable decrease of these infections in the general population [4,5].

Before the COVID-19 pandemic, an average of 48 hospitalizations with influenza per 100000 population were reported annually during 2008-2017 in Norway, with the highest incidence reported for those above 80 years [6]. While for RSV an average of 861 RSV hospitalizations per 100000 children under 5 years were reported annually in Norway [7]. RSV also represents a burden among elderly, with 39% of the annual RSV-associated hospitalizations in the European union (EU) being attributed to those aged 65 years and older [8].

In Norway, register-based surveillance of severe acute respiratory infections (SARI) was established in 2021 to prepare for the re-emergence of seasonal respiratory infections [9]. Describing the dynamics of respiratory infections may impact decisions on future protective strategies and immunization recommendations. Using data from the existing SARI surveillance, we assess the in-hospital disease burden of COVID-19, influenza, and RSV and investigate changes in the incidence and seasonality of hospitalizations with RSV and influenza in Norway after the emergence of COVID-19.

## Material and Methods

Utilizing the existing SARI-surveillance data, we assess the in-hospital disease burden, severity, length of stay (LOS) and seasonality of COVID-19, influenza, and RSV infections in Norway for the period 01 January 2017 to 30 April 2024.

### Data sources

The SARI surveillance system was established within the infrastructure of the Emergency preparedness register for COVID-19 (“Beredt C19”) at the Norwegian Institute of Public Health (https://www.fhi.no/en/id/corona/coronavirus/emergency-preparedness-register-for-covid-19/). Beredt C19 contained individual level data from national health registries, medical quality registries, and national administrative registries, covering the entire Norwegian population. Within the Beredt C19 framework, data on health region, diagnostic codes (ICD-10) and procedure codes (NCMP) were retrieved from The Norwegian Patient Registry (NPR), and laboratory test results were retrieved from the Norwegian Surveillance System for Communicable Diseases (MSIS) laboratory database. Sex and date of death were retrieved from the NPR and the National Population Register.

Between 2017 and August 2020, hospital admissions with influenza and RSV were defined using specific ICD-10 codes (Supplemental table 1). From September 2020, a positive PCR-test for SARS-CoV-2, influenza A, influenza B or RSV was also required. For admissions where no ICD-10 code was available, the case definition relied on laboratory results alone [9]. To account for transfers between wards and hospitals, we combined individual hospital stays within ≤2 days. Information on respiratory support and/or intensive care were identified using procedure codes (see Supplementary methods). Death was defined as any registered death between hospital admission until two weeks after discharge.

### Statistical analysis

We described the incidence and number of hospitalizations with COVID-19, influenza and RSV, between 01 January 2017 to 30 April 2024, by age group (0-4, 5-14, 15-29, 30-64, 65-79, ≥80 years and total), week and season (defined as a full year from week 26 until week 25 the following year). The yearly population per 1 January was retrieved from Statistics Norway and used as the denominator (https://www.ssb.no/).

We analyzed the 2022/2023 and 2023/2024 seasons to assess the LOS for discharged cases, and among hospitalized cases, proportions requiring intensive care and/or respiratory support and deaths while SARS-CoV-2, influenza, and RSV was co-circulating. The 2021/2022 season was excluded to avoid the immediate effects of potential immune debt. Using a Cox regression model, we estimated the hazard of discharge, stratified by age group, and adjusted for sex and health region. Odds of intensive care and/or respiratory support and death were compared using penalized maximum likelihood logistic regression, estimating odds ratios (OR) with 95% confidence intervals (CI) stratified by age group. Independent variables included infectious agent, sex and health region (with age groups for totals). Statistical analysis was performed in Stata version 18 (Stata Corporation, College Station, Texas, US).

### Ethical considerations

Ethical approval from the regional ethics committee was not required as the use of Beredt C19 was governed by The Act on Health and Social Preparedness §2-4, giving the Norwegian institute of public health a legal mandate to use registry data to implement and report on surveillance of COVID-19 and related infectious diseases. All analyses were conducted on pseudonymized data, and only anonymous aggregated data were exported from Beredt C19.

## Results

### Disease burden and re-establishment of seasonal epidemics, 2017-2024

Between 2017 and 2020, seasonal hospitalizations associated with influenza ranged from 3143 to 7737 (58.6–146.1 per 100000 population) and 834 to 2826 (15.5–53.0 per 100 000 population) for RSV (Table 1). The 2019–2020 influenza and RSV seasons ended early, coinciding with implementation of strict NIPs in March 2020 (Figure 1, Figure 2, Table 1). In 2020/2021, neither influenza nor RSV circulated in Norway (Figure 1, Figure 2).

**Figure 1:**
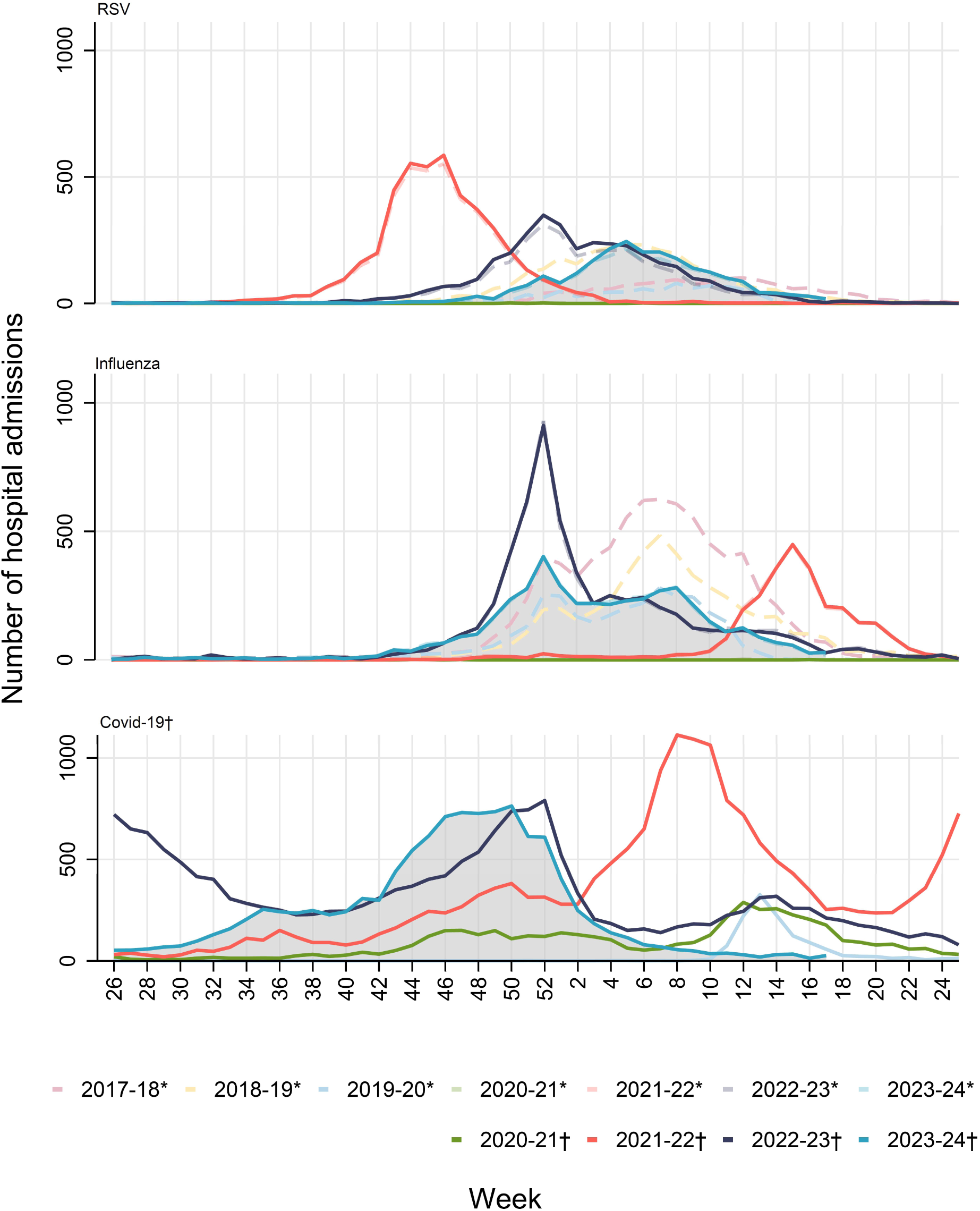
Number of weekly hospital admissions of respiratory syncytial virus (RSV), influenza and COVID-19 between week 26 and week 25 the following year, Norway, 26 June 2017 – 30 April 2024. *Admissions with an ICD-10 diagnosis. † Laboratory confirmed admissions.

**Figure 2:**
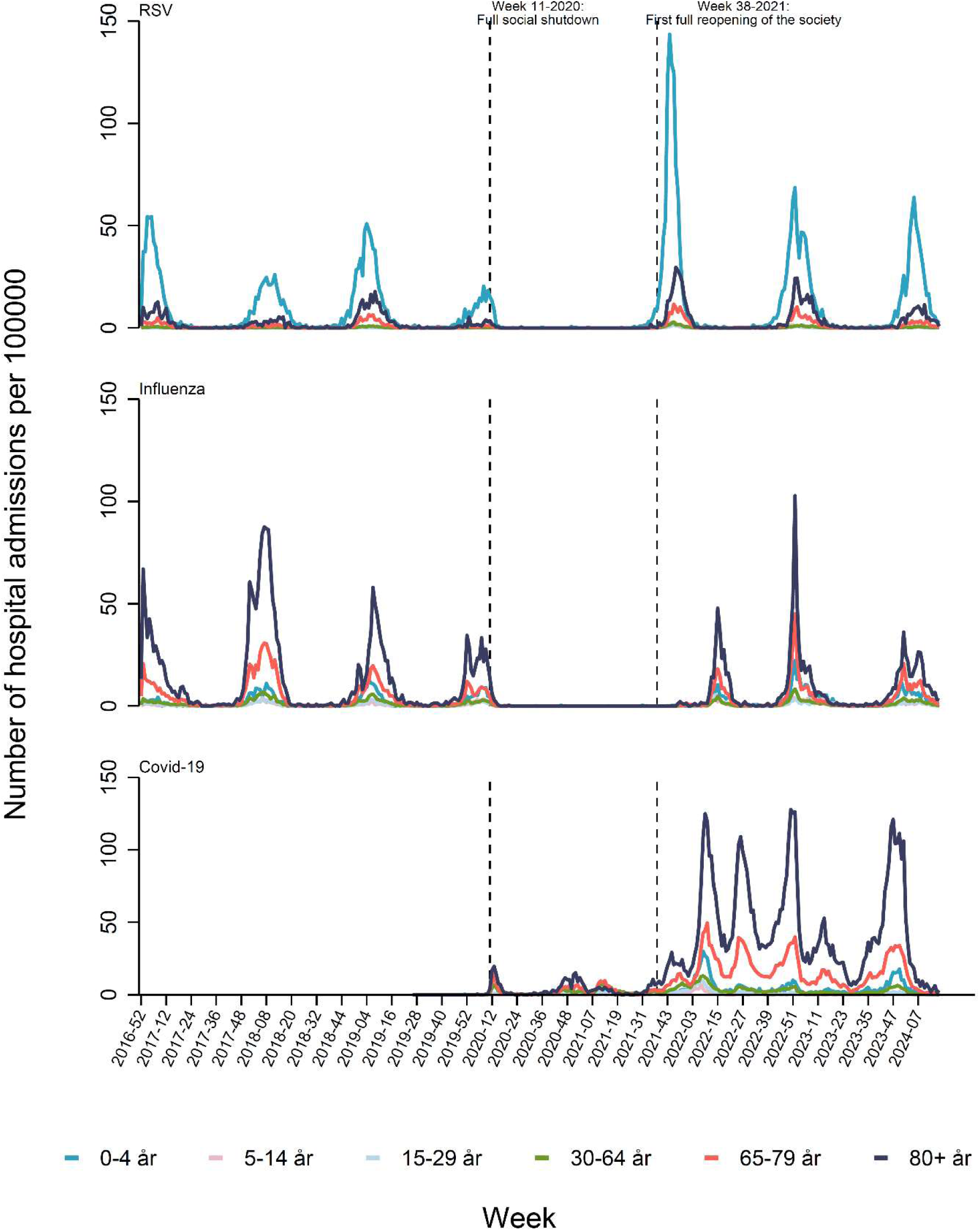
3-week moving average of the incidence of hospital admissions per 100000 population with respiratory syncytial virus (RSV), influenza and COVID-19 per age group, Norway 26 June 2017 – 30 April 2024.

**Table 1:** Total number (n) of hospitalized cases and incidence per 100000 population per year (week 26 to week 25 following year. 2024 was followed until week 17) for COVID-19, influenza and respiratory syncytial virus (RSV), Norway, 26 June 2017-30 April 2024.

| **Season** | **Covid-19**<br>**n (incidence per**<br>**100000 population)** | **RSV**<br>**n (incidence per**<br>**100000 population)** | **Influenza**<br>**n (incidence per**<br>**100000 population)** |
| --- | --- | --- | --- |
| 2017/2018* | Not applicable | 1456 (27.5) | 7737 (146.1) |
| 2018/2019* | Not applicable | 2826 (53.0) | 4891 (91.8) |
| 2019/2020* | 1255 (23.4) | 834 (15.5) | 3143 (58.6) |
| 2020/2021 | 4635 (86.0) | 9 (0.2) | 5 (0.1) |
| 2021/2022 | 17421 (321.1) | 4498 (82.9) | 2810 (51.8) |
| 2022/2023 | 17131 (312.1) | 3563 (64.9) | 5911 (107.7) |
| 2023/2024† | 11000 (200.4) | 2368 (43.1) | 4406 (80.3) |

\* Number of diagnoses is based on ICD-10 codes only, without laboratory confirmation
† For season 2023 - 2024 data was only available until 30 April 2024.

As measures against COVID-19 were gradually eased towards week 38, 2021, RSV associated hospitalizations surged, leading to an unusually early epidemic, that lasted until January 2022 (Figure 1, Figure 2 and Supplemental Figure 2). RSV hospitalization incidence during this season were ∼56% higher than any pre-pandemic season (Table 1), with children under 5 years old experiencing the highest rates (946 RSV hospitalizations per 100 000, Supplemental table 2). Meanwhile, COVID-19 hospitalizations rose gradually in autumn 2021, culminating in a major wave beginning in January 2022 as the RSV epidemic subsided. COVID-19 remained the leading cause of hospitalizations from respiratory infections until April 2022.

Influenza circulation was minimal through late 2021, with ≤1% of SARI hospitalizations linked to influenza by the end of the year. A delayed influenza epidemic began in March 2022 lasting until in early June before a second wave of COVID-19 emerged, lasting until late August 2022 (Figure 1, Figure 2, Supplemental Figures 2 and 3).

By autumn 2022, influenza and RSV hospitalization trends aligned more closely with pre-pandemic timing. Both viruses peaked alongside a COVID-19 wave in late December 2022, resulting in a “tripledemic” (Figure 1). The RSV and influenza epidemics ended by week 16, 2023, while COVID-19 hospitalizations persisted through the season, reaching a low in week 26 before peaking again in week 50, 2023. In autumn 2023, influenza hospitalizations rose in week 42, peaking in week 52, while RSV peaked later in week 5 of 2024.

During the three seasons of SARS-CoV-2, influenza virus and RSV co-circulation, COVID-19 hospitalizations outnumbered influenza hospitalizations by 3.5 times (range: 2.5–6.2) and RSV hospitalizations by 4.4 times (range: 3.9–4.8) on average (Table 1). Those ≥65 years accounted for the majority of COVID-19 and influenza hospitalizations (∼67% and 52%, respectively), whereas RSV hospitalizations were more common among children under 5 (∼58%) and less frequent in those ≥65 (∼29%, Supplemental table 2).

Length of stay, 2022-2024

Younger age groups had a shorter LOS compared to older age groups for all three diseases. A small number of patients had very long LOS, and a LOS >90 days were treated as outliers (values including outliers are shown in brackets when the difference is ≥0.5 days). For COVID-19 admissions, patients <30 years old had an average LOS of ≤4.6 days (≤5.9 days including outliers), while those aged ≥30 years had an average LOS of ≥6.1 days. A similar pattern was observed for influenza admissions with ≤3.1 days (≤3.6 days with outliers) for patients under 30 and ≥4.4 days for those 30 and older, and for RSV admissions with ≤4.6 days for patients under 30 and ≥6.3 days for those 30 and older (Table 2, Supplemental table 3).

**Table 2:**
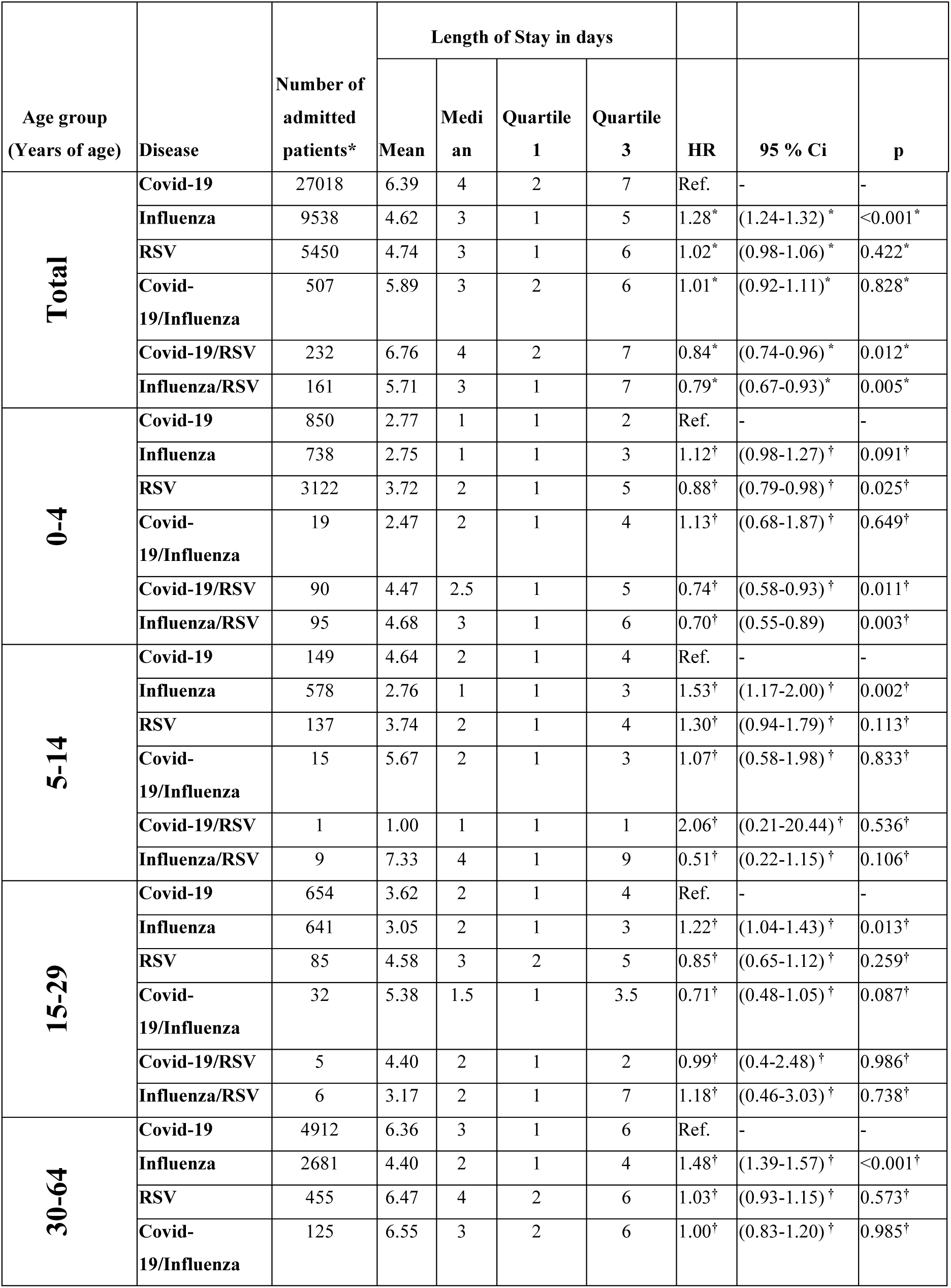
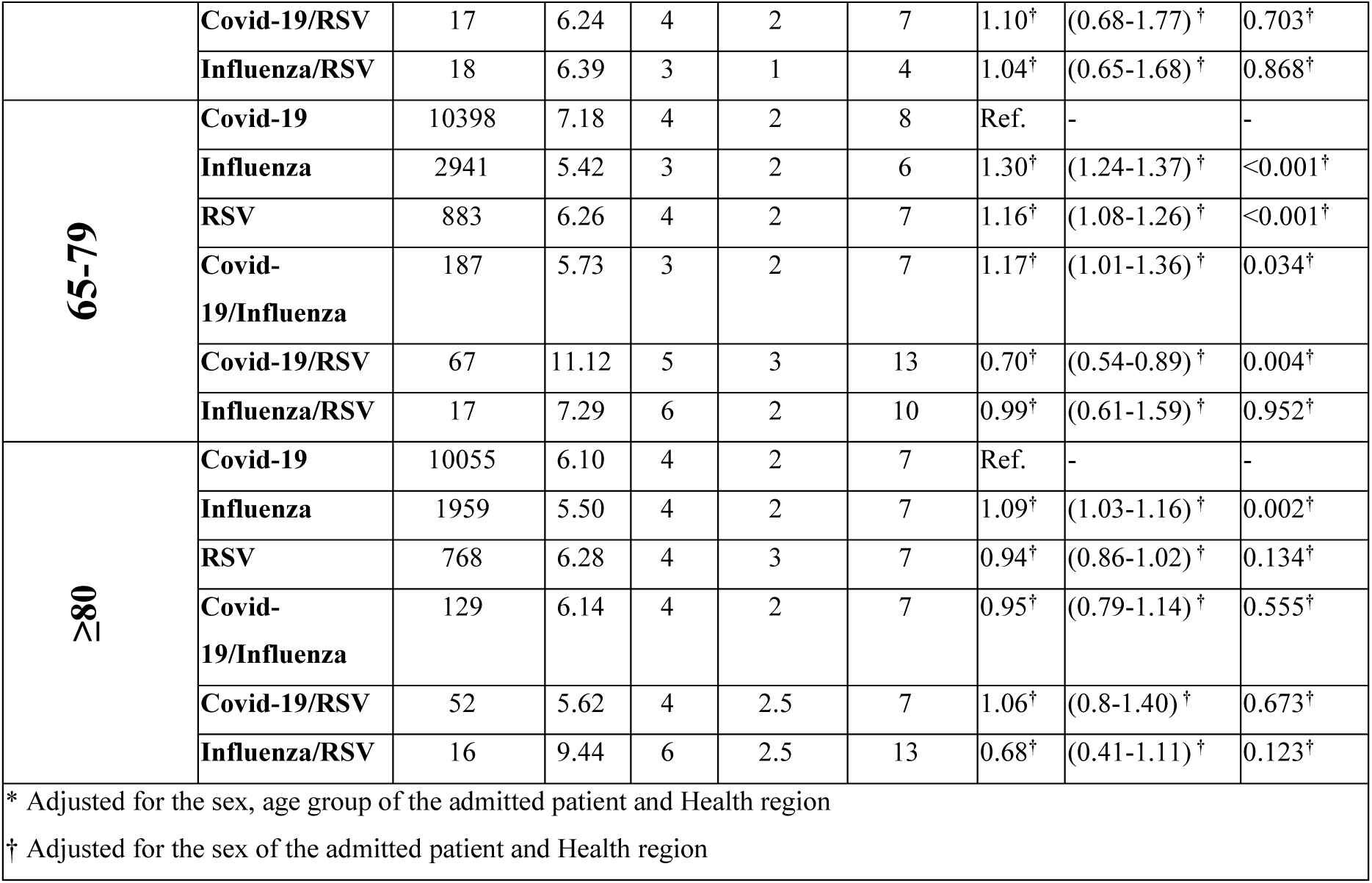
Length of stay in days and hazard ratio (HR) for discharge per age group for patients admitted with COVID-19, influenza and respiratory syncytial virus (RSV) in Norway between 27 June 2022 – 30 April 2024. Excluding outliers with a stay of more than 90 days

| Age group<br>(Years of age) | Disease | Number of<br>admitted<br>patients* | Length of Stay in days |  |  |  | HR | 95 % Ci | p |
| --- | --- | --- | --- | --- | --- | --- | --- | --- | --- |
|  |  |  | Mean | Median | Quartile<br>1 | Quartile<br>3 |  |  |  |
| **Total** | Covid-19 | 27018 | 6.39 | 4 | 2 | 7 | Ref. | - | - |
|  | Influenza | 9538 | 4.62 | 3 | 1 | 5 | 1.28* | (1.24-1.32)* | <0.001* |
|  | RSV | 5450 | 4.74 | 3 | 1 | 6 | 1.02* | (0.98-1.06)* | 0.422* |
|  | Covid-19/Influenza | 507 | 5.89 | 3 | 2 | 6 | 1.01* | (0.92-1.11)* | 0.828* |
|  | Covid-19/RSV | 232 | 6.76 | 4 | 2 | 7 | 0.84* | (0.74-0.96)* | 0.012* |
|  | Influenza/RSV | 161 | 5.71 | 3 | 1 | 7 | 0.79* | (0.67-0.93)* | 0.005* |
| **0-4** | Covid-19 | 850 | 2.77 | 1 | 1 | 2 | Ref. | - | - |
|  | Influenza | 738 | 2.75 | 1 | 1 | 3 | 1.12† | (0.98-1.27)† | 0.091† |
|  | RSV | 3122 | 3.72 | 2 | 1 | 5 | 0.88† | (0.79-0.98)† | 0.025† |
|  | Covid-19/Influenza | 19 | 2.47 | 2 | 1 | 4 | 1.13† | (0.68-1.87)† | 0.649† |
|  | Covid-19/RSV | 90 | 4.47 | 2.5 | 1 | 5 | 0.74† | (0.58-0.93)† | 0.011† |
|  | Influenza/RSV | 95 | 4.68 | 3 | 1 | 6 | 0.70† | (0.55-0.89) | 0.003† |
| **5-14** | Covid-19 | 149 | 4.64 | 2 | 1 | 4 | Ref. | - | - |
|  | Influenza | 578 | 2.76 | 1 | 1 | 3 | 1.53† | (1.17-2.00)† | 0.002† |
|  | RSV | 137 | 3.74 | 2 | 1 | 4 | 1.30† | (0.94-1.79)† | 0.113† |
|  | Covid-19/Influenza | 15 | 5.67 | 2 | 1 | 3 | 1.07† | (0.58-1.98)† | 0.833† |
|  | Covid-19/RSV | 1 | 1.00 | 1 | 1 | 1 | 2.06† | (0.21-20.44)† | 0.536† |
|  | Influenza/RSV | 9 | 7.33 | 4 | 1 | 9 | 0.51† | (0.22-1.15)† | 0.106† |
| **15-29** | Covid-19 | 654 | 3.62 | 2 | 1 | 4 | Ref. | - | - |
|  | Influenza | 641 | 3.05 | 2 | 1 | 3 | 1.22† | (1.04-1.43)† | 0.013† |
|  | RSV | 85 | 4.58 | 3 | 2 | 5 | 0.85† | (0.65-1.12)† | 0.259† |
|  | Covid-19/Influenza | 32 | 5.38 | 1.5 | 1 | 3.5 | 0.71† | (0.48-1.05)† | 0.087† |
|  | Covid-19/RSV | 5 | 4.40 | 2 | 1 | 2 | 0.99† | (0.4-2.48)† | 0.986† |
|  | Influenza/RSV | 6 | 3.17 | 2 | 1 | 7 | 1.18† | (0.46-3.03)† | 0.738† |
| **30-64** | Covid-19 | 4912 | 6.36 | 3 | 1 | 6 | Ref. | - | - |
|  | Influenza | 2681 | 4.40 | 2 | 1 | 4 | 1.48† | (1.39-1.57)† | <0.001† |
|  | RSV | 455 | 6.47 | 4 | 2 | 6 | 1.03† | (0.93-1.15)† | 0.573† |
|  | Covid-19/Influenza | 125 | 6.55 | 3 | 2 | 6 | 1.00† | (0.83-1.20)† | 0.985† |
|  | **Covid-19/RSV** | 17 | 6.24 | 4 | 2 | 7 | 1.10<sup>†</sup> | (0.68-1.77)<sup>†</sup> | 0.703<sup>†</sup> |
|  | **Influenza/RSV** | 18 | 6.39 | 3 | 1 | 4 | 1.04<sup>†</sup> | (0.65-1.68)<sup>†</sup> | 0.868<sup>†</sup> |
| **65-79** | **Covid-19** | 10398 | 7.18 | 4 | 2 | 8 | Ref. | - | - |
|  | **Influenza** | 2941 | 5.42 | 3 | 2 | 6 | 1.30<sup>†</sup> | (1.24-1.37)<sup>†</sup> | <0.001<sup>†</sup> |
|  | **RSV** | 883 | 6.26 | 4 | 2 | 7 | 1.16<sup>†</sup> | (1.08-1.26)<sup>†</sup> | <0.001<sup>†</sup> |
|  | **Covid-19/Influenza** | 187 | 5.73 | 3 | 2 | 7 | 1.17<sup>†</sup> | (1.01-1.36)<sup>†</sup> | 0.034<sup>†</sup> |
|  | **Covid-19/RSV** | 67 | 11.12 | 5 | 3 | 13 | 0.70<sup>†</sup> | (0.54-0.89)<sup>†</sup> | 0.004<sup>†</sup> |
|  | **Influenza/RSV** | 17 | 7.29 | 6 | 2 | 10 | 0.99<sup>†</sup> | (0.61-1.59)<sup>†</sup> | 0.952<sup>†</sup> |
| **≥80** | **Covid-19** | 10055 | 6.10 | 4 | 2 | 7 | Ref. | - | - |
|  | **Influenza** | 1959 | 5.50 | 4 | 2 | 7 | 1.09<sup>†</sup> | (1.03-1.16)<sup>†</sup> | 0.002<sup>†</sup> |
|  | **RSV** | 768 | 6.28 | 4 | 3 | 7 | 0.94<sup>†</sup> | (0.86-1.02)<sup>†</sup> | 0.134<sup>†</sup> |
|  | **Covid-19/Influenza** | 129 | 6.14 | 4 | 2 | 7 | 0.95<sup>†</sup> | (0.79-1.14)<sup>†</sup> | 0.555<sup>†</sup> |
|  | **Covid-19/RSV** | 52 | 5.62 | 4 | 2.5 | 7 | 1.06<sup>†</sup> | (0.8-1.40)<sup>†</sup> | 0.673<sup>†</sup> |
|  | **Influenza/RSV** | 16 | 9.44 | 6 | 2.5 | 13 | 0.68<sup>†</sup> | (0.41-1.11)<sup>†</sup> | 0.123<sup>†</sup> |
| * Adjusted for the sex, age group of the admitted patient and Health region |  |  |  |  |  |  |  |  |  |
| † Adjusted for the sex of the admitted patient and Health region |  |  |  |  |  |  |  |  |  |

Influenza admissions generally resulted in slightly shorter LOS than COVID-19 across all age groups, except for children aged 0–4 years. RSV admissions were associated with longer stays in the 0–4 age group but shorter stays in the 65–79 age group compared to COVID-19 admissions (Table 2).

### Ventilatory support, intensive care, and deaths, 2022-2024

Between 2022-2024, ventilatory support and/or intensive care was registered for 7.9% of COVID-19 admissions, 7.8% of influenza admissions and 18.0% of RSV admissions. There was no differences in the proportion receiving ventilatory support and/or intensive care between COVID-19 and influenza hospitalizations, except in the 5-14 year olds (Table 3, Supplemental table 4).

**Table 3:**
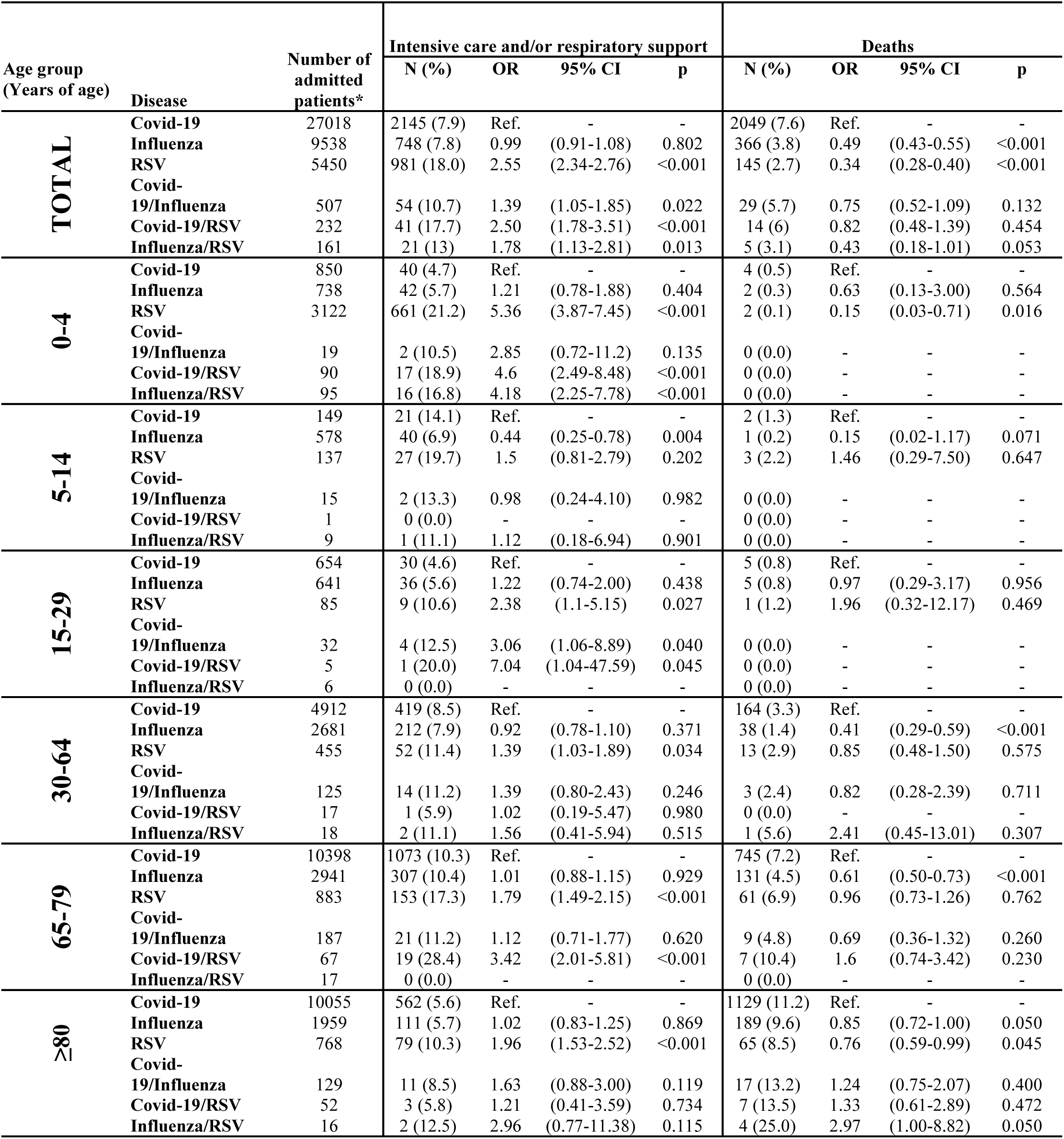
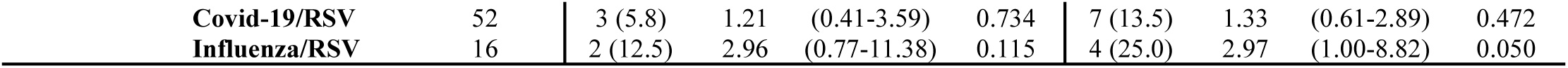
Comparison of use of intensive care and/or ventilatory support (invasive mechanical ventilation, continuous positive airway pressure [CPAP] or bi-level positive airway pressure [BiPAP]) during the hospital stay, and number of deaths occurred during admission or within two weeks of discharge among patients admitted with COVID-19, influenza or respiratory syncytial virus (RSV) in Norway, between 27 June 2022 – 30 April 2024. Excluding outliers with a stay of more than 90 days.

In contrast, ventilatory support and/or intensive care was used more frequently for RSV hospitalizations compared to COVID-19 hospitalizations across most age groups (except 5-14 years old). The largest difference was observed in children aged 0–4 years, where 21% of RSV admissions and 5% of COVID-19 admissions received ventilatory support and/or intensive care (OR [95% CI]: 5.4 [3.87–7.45]).

Patients with co-infections (SARS-CoV-2 + influenza, SARS-CoV-2 + RSV, or influenza + RSV) were also more likely to receive ventilatory support and/or intensive care compared to those testing positive for only SARS-CoV-2 (Table 3, Supplemental table 4).

Deaths within two weeks of hospitalization were rare among patients <30 years of age. The percentage of reported deaths increased with age and was highest in the age group 80 years or older for all three diseases (Table 3). Overall, the percentage of COVID-19 hospitalizations associated with death (7.6%) was twice that of influenza hospitalizations (3.8%) and almost three times that of RSV hospitalizations (2.7%).

## Discussion

During the first year of the COVID-19 pandemic, the transmission of influenza and RSV was absent. From 2021 to 2024, COVID-19 accounted for a larger share of SARI-hospitalizations compared to influenza and RSV and had a higher mortality rate within two weeks of admission. However, patients hospitalized with RSV were more likely to require ventilatory support or intensive care.

### Disease burden and re-establishment of seasonal epidemics

In Norway, the first COVID-19 case was detected on February 26, 2020 [10]. Following a rapid increase in COVID-19 hospitalizations, the Norwegian government implemented a national lockdown in mid-March, accompanied by strict infection control measures. While these measures gradually eased towards the summer 2020, restrictions remained in place and varied by region, depending on the local COVID-19 epidemiological situation [11]. Most restrictions were lifted on September 25, 2021 [12]. During the 2020-2021 period, hospitalizations for influenza and RSV were virtually nonexistent in Norway. We attribute the absence of influenza and RSV primarily to behavioral changes driven by the evolving COVID-19 crisis and in response to NPIs and heightened health risk perceptions.

Many countries reported similar patterns of absent influenza and RSV epidemics during the COVID-19 pandemic, followed by uncharacteristic outbreaks outside the usual seasonal timeframe once COVID-19 restrictions were eased [4, 5, 13]. The similar timing and age distribution of RSV outbreaks in several countries during the summer and autumn of 2021 suggest that immunity debt, resulting from the shortened 2019/2020 season and the absence of cases in 2020/2021, was a key contributing factor [14]. One could speculate on the role of heterologous viral interference between SARS-CoV-2, influenza virus and RSV in shaping of the unusual seasonal patterns observed as influenza and RSV circulation resumed [15]. However, the “tripledemic” during the season of 2022/23 demonstrated the potential for SARS-CoV-2, influenza virus and RSV to co-circulate. It is likely that multiple factors contributed to the re-emergence of respiratory diseases. In the case of influenza, sporadic cases were seen starting in November 2021, coinciding with a gradual rise in COVID-19 cases driven by the Delta variant [16]. By the end of November 2021, NIPs were again implemented [17]. lasting until February 12, 2022 [18], which likely delayed the influenza season.

There is a clear age difference in the distribution of hospitalizations with COVID-19, influenza and RSV. Between 2022 and 2024, most patients hospitalized with COVID-19, and just over half of those hospitalized with influenza were aged 65 years or older. In contrast, less than one-third of RSV patients fell into this age group. The vaccine recommendations for COVID-19 and influenza were similar. However, the vaccine coverage among individuals aged 65 and older was about 54% against COVID-1919 and 64% for influenza in 2023/2024 [19, 20]. Across all age groups, coverage was 12% for COVID-19 and 23% for influenza (only counting vaccinations from September 2023) [19]. This difference in vaccine coverage may have contributed to the higher number of hospitalizations with COVID-19. It should be noted that both the 2022/2023 and 2023/2024 seasons were dominated by influenza A, with some circulation of influenza B towards the end of the epidemics. During the 2022/2023 season there was a higher proportion of children among those hospitalized with influenza B compared with influenza A [21]. However, we could not observe any obvious shift in the age distribution compared to the years before the emergence of SARS-CoV-2 (Figure 2). Our findings show a higher incidence of influenza hospitalization than previously reported in Norway. Hauge et al. found an incidence of 62 per 100000 among individuals aged 60-69 and 241 per 100000 for those over 80 years. However, while Hauge et al. excluded repeat patients within a 120-day period to reflect the incidence of influenza cases in the population [6], our study focused on the incidence of new hospitalizations.

The age distribution of RSV hospitalizations remained similar before and after the COVID-19 pandemic. Before the COVID-19 pandemic, individuals aged 65 years and older accounted for 39% of annual RSV-associated hospitalizations in the EU [8], while during the 2022/2023 and 2023/2024 seasons, 31% of RSV hospitalizations were in this age group. Similarly, ∼57% of RSV cases were children <5 years old, compared to ∼60% before the COVID-19 pandemic [7, 8]. The incidence of the 2022/2023 and 2023/2024 seasons is comparable to estimates from a previous large seasonal epidemic (2016/2017) in Norway for children <5 years [22], with an even higher incidence during the 2021/2022 peak. The typical pattern of biennial low and high seasons [23] has not yet been re-established.

### Length of stay, ventilatory support, intensive care, and deaths

Since immunization measures for RSV differ significantly from those for influenza and COVID-19 – and are likely to change – the results of this study should not guide future assessments of disease severity. Instead, they provide a comparison of the hospital burden these diseases caused in Norway during the study period. Most studies comparing disease severity between COVID-19, influenza and RSV patients, report longer LOS and higher mortality among COVID-19 patients than among influenza patients [24–26]. These studies often compare COVID-19 data from the pandemic’s first two years with historical influenza cohorts [24, 25], or from 2020, when influenza circulation was greatly reduced [26]. In this study, we compared co-circulating COVID-19, influenza and RSV, finding higher mortality among elderly COVID-19 patients compared to influenza. Notably, the longer LOS and higher mortality associated with COVID-19 did not lead to greater demand for intensive care or respiratory support. A study on Omicron-dominant periods found no differences in LOS, mortality, intensive care admissions, or respiratory support between COVID-19 and influenza patients [27]. While our results align partly with Omicron-era findings [27], they also partly reflect trends from early-pandemic studies [24–26], showing higher mortality and longer LOS for COVID-19 but no difference in intensive care and/or need of respiratory support needs. This may reflect greater frailty among COVID-19 fatalities compared to those from influenza [28].

COVID-19 has also been associated with longer intensive care unit stays and higher mortality compared to RSV in adults [29]. Overall, we found no significant difference in LOS between COVID-19 and RSV, though RSV patients under 5 years old had longer stays. Similarly, another study has reported longer LOS for RSV than COVID-19 among hospitalized children <3-year-old [30].

RSV hospitalizations had a higher risk of requiring intensive care and/or ventilatory support compared to COVID-19 hospitalizations. A recent study from Norway found that <50% of SARI cases above 65 years were tested for RSV, while 82% were tested for SARS-CoV-2 [9]. This testing disparity could impact the proportion of intensive care admissions among RSV cases, as limited RSV testing may skew results toward more severe cases. Consequently, milder RSV cases may be underrepresented in RSV-specific ICD-10 codes, leading to an overestimation of intensive care and/or ventilatory support needs. However, since 80% of SARI cases in children aged 0–4 were tested for RSV and 87% for SARS-CoV-2 [9], testing differences cannot fully explain the higher proportion of RSV cases requiring intensive care or ventilatory support in this youngest age group.

### Strengths and weaknesses

This study does not account for protective measures such as vaccination, monoclonal antibodies, antiviral treatments, or NPIs (e.g., masking or social distancing). As noted, the vaccination coverage varied between influenza and COVD-19 [19, 20], potentially influencing hospitalization rates. No data of RSV-vaccination coverage were available at the time of writing.

A strength of this study is the use of national health registry data linked to the national laboratory database, enabling analysis of laboratory-confirmed cases and reducing reliance on ICD-10 codes, which underreport RSV incidence [9]. However, as the laboratory database only became available in 2020, different case definitions were used before the COVID-19 pandemic. Despite this, the close correlation of identified cases using these two methods (Supplemental table 5), suggests this is a minor issue, though RSV incidence in pre-pandemic seasons may be slightly underestimated.

## Conclusion

Influenza and RSV transmission was disrupted during the first year of the COVID-19 pandemic, followed by unusual seasonality in the year after societal reopening. RSV has yet to reestablish pre-pandemic circulation patterns, with all post-pandemic seasons resembling high pre-pandemic RSV seasons. While RSV and influenza significantly contributed to hospital burden, COVID-19 was the largest contributor during the initial years of co-circulation.

## List of abbreviations

BeredtC19: Norwegian national preparedness registry for COVID-19
aHR: Adjusted hazard ratio
ALRI: Acute lower respiratory infections
CI: Confidence intervals
KUHR: Norwegian Control and Payment of Health Reimbursements Database
LOS: Length of stay
NPIs: Non-pharmaceutical interventions
NPR: The Norwegian Patient Registry
OR: Odds ratio
RSV: Respiratory syncytial virus
SARI: Severe Acute Respiratory Infections

## Declarations

### Availability of data and materials

The datasets analyzed during the current study come from the national emergency preparedness registry for COVID-19, housed at the Norwegian Institute of Public Health. The preparedness registry comprised data from a variety of central health registries, national clinical registries and other national administrative registries. Further information on the preparedness registry, including access to data from each data source, is available at https://www.fhi.no/en/id/infectious-diseases/coronavirus/emergency-preparedness-register-for-COVID-19/.

### Authors’ contributions

All co-authors were involved in the conceptualization of the study. HB, ES, LV and THP drafted the study protocol and coordinated the study. HB, ES, THP and JS contributed directly to the acquisition of data. HB, ES and JD contributed to data cleaning, verification and preparation. HB, ES, JS and JD had access to the final linked dataset. HB conducted the statistical analysis with support from ES. All co-authors contributed to the interpretation of the results. HB and LV drafted the manuscript. All co-authors contributed to the revision of the manuscript and approved the final version for submission.

## Data Availability

The datasets analysed during the current study come from the national emergency preparedness registry for COVID-19. The preparedness registry comprised data from a variety of central health registries, national clinical registries and other national administrative registries. Further information on the preparedness registry is available at https://www.fhi.no/en/id/corona/coronavirus/emergency-preparedness-register-for-covid-19/. Access to data from each individual data source is application based through https://helsedata.no/en/, after approval from the Norwegian Committee for Medical and Health Research Ethics and within the framework of the Norwegian data protection legislation.

