## Supplemental material for "Changed epidemiology of influenza and RSV hospitalizations after the emergence of SARS-CoV-2 in Norway, 2017 - 2024"

*Supplementary methods*

Information on respiratory support was retrieved using the Norwegian procedural codes GXAV01 – invasive mechanical ventilation, GXAV10 - Continuous positive airway pressure (CPAP), GXAV20 - Bilevel positive airway pressure (BiPAP) while information on intensive care admissions were identified using the Norwegian procedural code B0050, which was in use from January 2022.

*Supplementary results*

*Comparison of RSV and influenza case definition using pathogen specific ICD-10 codes or SARI codes in addition to laboratory conformation*

Weekly number of hospitalizations identified using ICD-10 codes (Supplemental table 1) specific for each of COVID-19, Influenza and RSV, correlated highly with cases identified using wider SARI ICD-10 codes in combination with laboratory confirmation (Figure 1 and Supplemental figure 1-3). Since 2020, SARI ICD-10 codes in combination with laboratory confirmation of Influenza identified 123 fewer cases than when relying on Influenza specific ICD-10 codes alone (Pearson correlation: 0.999, P-value <0.001, Supplemental table 5), with no major deviations within the individual age groups.

SARI ICD-10 codes in combination with laboratory confirmation of RSV, identified more RSV cases compared to RSV specific codes without laboratory confirmation (900 extra RSV cases, Pearson correlation: 0.999, P-value <0.001). However, the deviation was spread out between the age groups, with the largest percent increase among 5-14- and 15–29-year-olds (58.6% and 57.3% respectively), and the highest number of additional cases among children <5 years (253 cases), followed by the 65–79-year-olds (218 cases, Supplemental table 5).

*Sup. Figures*

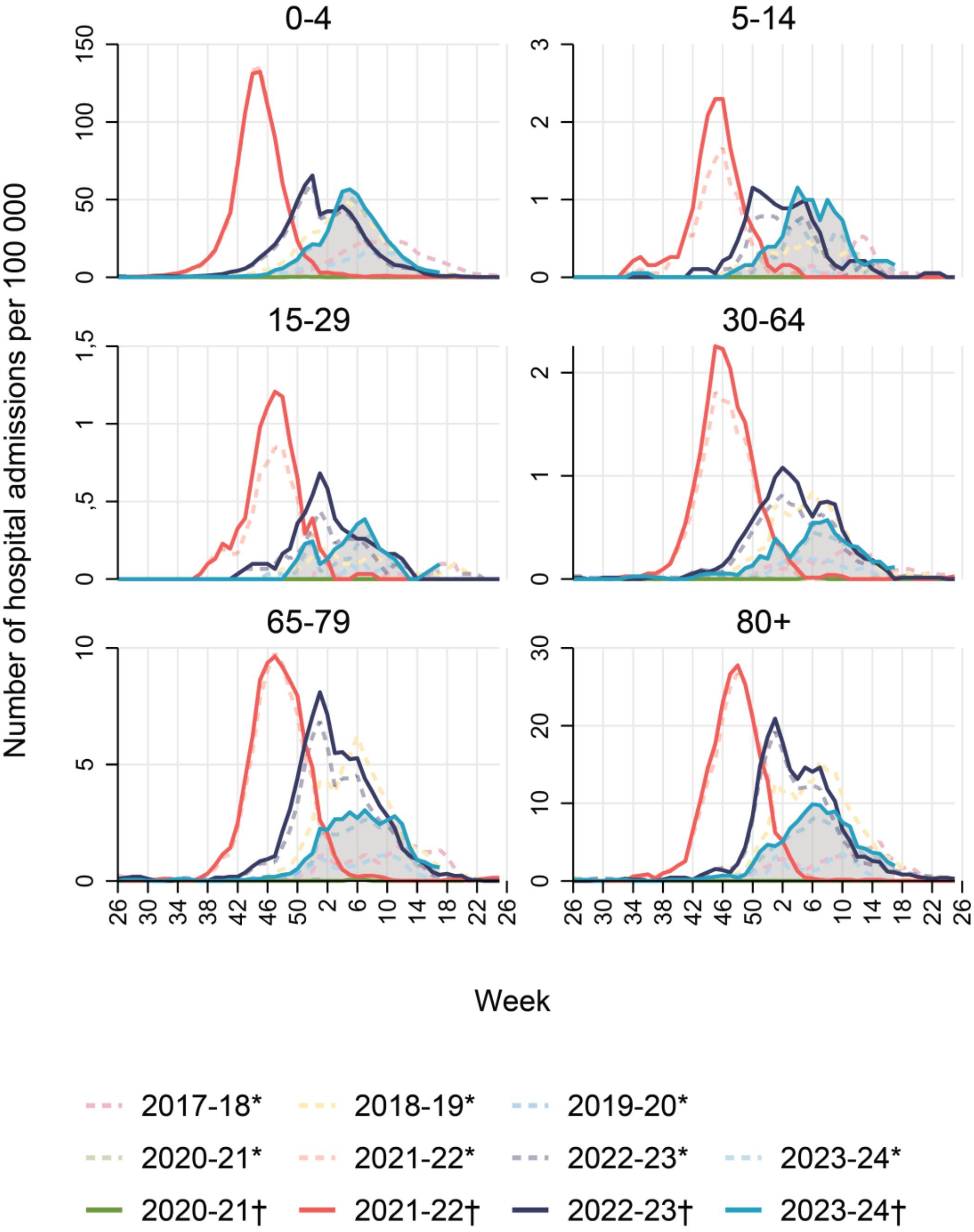


**Supplemental figure 1: Respiratory syncytial virus (RSV) incidence by age group and year, using three weeks moving average (per 100000 population) of hospital admissions between week 26 and week 25 the following year, Norway, 26 June 2017 – 30 April 2024. *Admissions with an ICD-10 diagnosis. † Laboratory confirmed admissions.**

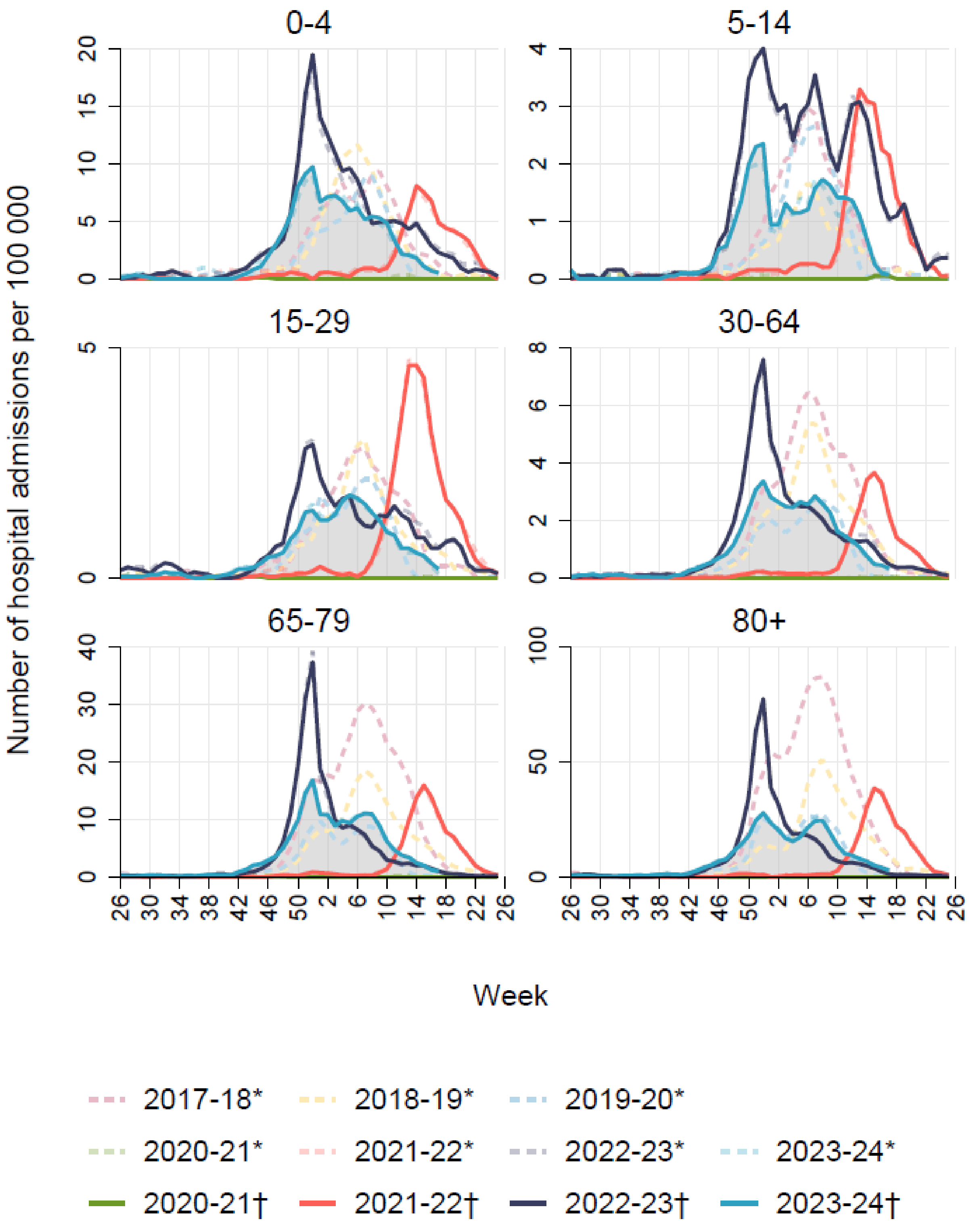


**Supplemental figure 2: Influenza incidence by age group and year, using three weeks moving average (per 100000 population) of hospital admissions between week 26 and week 25 the following year, Norway, 26 June 2017 – 30 April 2024. *Admissions with an ICD-10 diagnosis. † Laboratory confirmed admissions.**

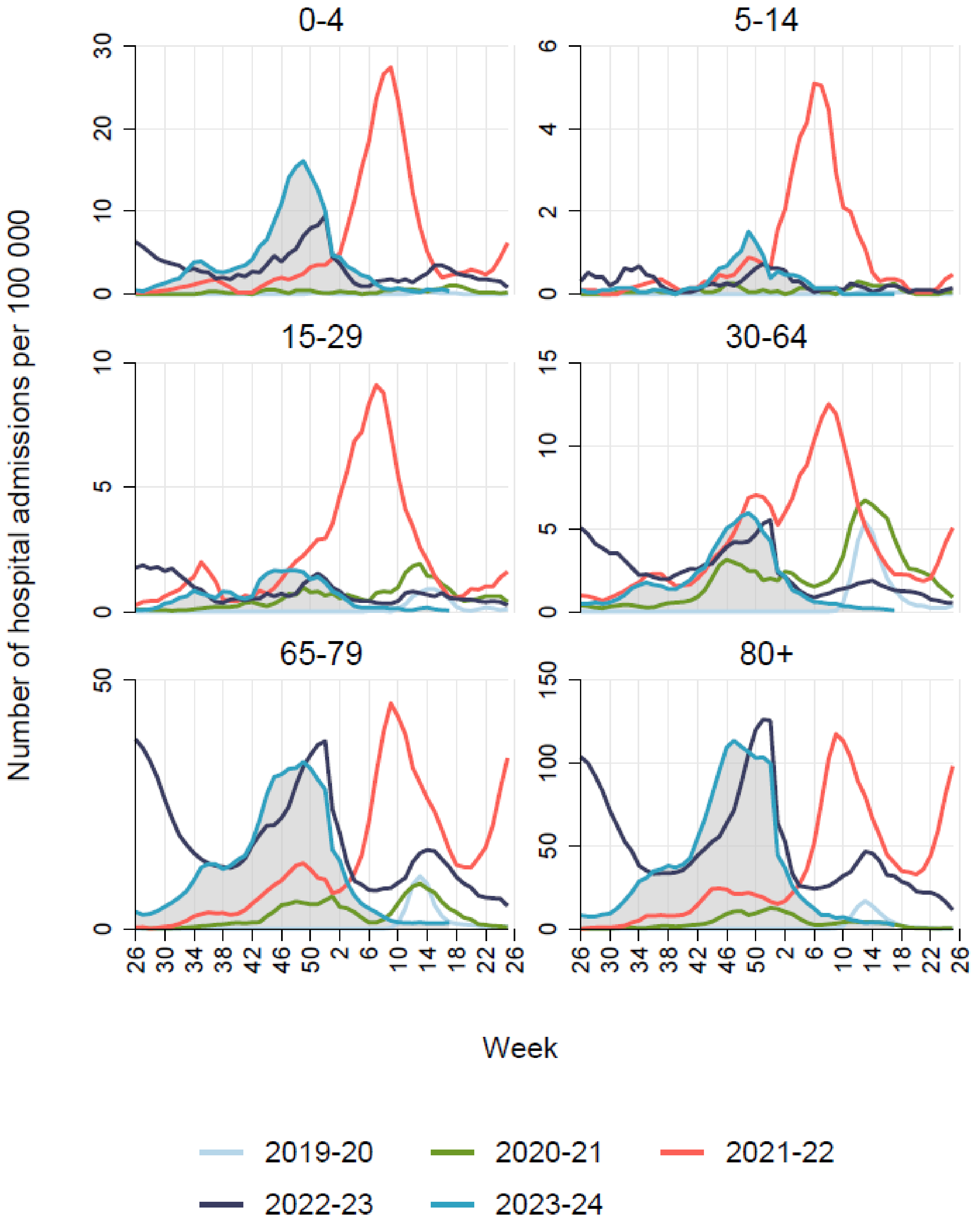


**Supplemental figure 3: COVID-19 incidence by age group and year, using three weeks moving average (per 100000 population) of hospital admissions between week 26 and week 25 the following year, Norway, 26 June 2017 – 30 April 2024. *Admissions with an ICD-10 diagnosis. † Laboratory confirmed admissions.**

*Sup. Tables*

**Supplemental table 1: ICD-10 codes used for SARI surveillance, Norway, 2017-2024.**

| **ICD-10 code** | **Description** |
|---|---|
| J00-J06 | Acute upper respiratory infections |
| J09-J22 | Influenza and pneumonia,<br>Other acute lower respiratory infections |
| J09-J11 | Influenza |
| J12.1, J20.5 and J21.0 | Respiratory Syncytial Virus Infection |
| J80 | Acute respiratory distress syndrome in adults |
| U07.1, U07.2 | COVID-19 |
| A37 | Whooping cough due to Bordetella pertussis |
| H65.0, H65.1, H65.9, H66.0, H66.4, H66.9 and H67 | Otitis media (acute and unspecified only) |

**Supplemental table 2: Total number of hospitalized cases and incidence per year (week 26 to week 25 following year) and age group for each of the diseases COVID-19, influenza and respiratory syncytial virus (RSV), Norway, 26 June 2017 – 30 April 2024.**

| | Age groups n (incidence per 100000 population) | | | | | |
|---|---|---|---|---|---|---|
| **Season** | **0-4** | **5-14** | **15-29** | **30-64** | **65-79** | **≥80** |
| | **COVID-19** | | | | | |
| **2019/2020** | 5 (1.72) | 5 (0.78) | 59 (5.73) | 660 (26.77) | 343 (48.23) | 183 (79.32) |
| **2020/2021** | 45 (15.90) | 30 (4.70) | 308 (30.15) | 2737 (110.25) | 1067 (146.30) | 448 (189.5) |
| **2021/2022** | 774 (276.39) | 319 (50.16) | 1245 (122.02) | 5561 (222.49) | 5260 (702.10) | 4262 (1773.67) |
| **2022/2023** | 469 (168.84) | 90 (14.10) | 421 (40.97) | 3113 (122.85) | 6777 (885.77) | 6261 (2541.4) |
| **2023/2024**† | 504 (181.44) | 79 (12.38) | 278 (27.06) | 2029 (80.07) | 4043 (528.43) | 4067 (1650.84) |
| | **Influenza** | | | | | |
| **2017/2018*** | 354 (118.17) | 195 (65.09) | 383 (127.85) | 1890 (630.89) | 2570 (857.87) | 2345 (782.77) |
| **2018/2019*** | 345 (117.00) | 110 (37.31) | 329 (111.58) | 1455 (493.45) | 1466 (497.18) | 1186 (402.22) |
| **2019/2020*** | 253 (87.22) | 166 (57.23) | 255 (87.91) | 830 (286.14) | 840 (289.59) | 799 (275.46) |
| **2020/2021** | 1 (0.35) | 1 (0.16) | 1 (0.10) | 0 (0.00) | 2 (0.27) | 0 (0) |
| **2021/2022** | 191 (68.20) | 159 (25.00) | 361 (35.38) | 639 (25.57) | 819 (109.32) | 641 (266.76) |
| **2022/2023** | 536 (192.95) | 428 (67.06) | 412 (40.10) | 1646 (64.96) | 1729 (225.98) | 1160 (470.86) |
| **2023/2024†** | 323 (116.28) | 183 (28.67) | 273 (26.57) | 1200 (47.36) | 1462 (191.09) | 965 (391.7) |
| | **RSV** | | | | | |
| **2017/2018*** | 1035 (345.48) | 20 (6.68) | 8 (2.67) | 92 (30.71) | 158 (52.74) | 143 (47.73) |
| **2018/2019*** | 1651 (559.92) | 33 (11.19) | 25 (8.48) | 235 (79.70) | 455 (154.31) | 427 (144.81) |

| | | | | | | |
|---|---|---|---|---|---|---|
| **2019/2020*** | 570 (196.51) | 10 (3.45) | 9 (3.10) | 62 (21.37) | 100 (34.48) | 83 (28.61) |
| **2020/2021** | 5 (1.77) | 0 (0.00) | 0 (0.00) | 1 (0.04) | 2 (0.27) | 1 (0.42) |
| **2021/2022** | 2648 (945.58) | 107 (16.83) | 94 (9.21) | 459 (18.36) | 651 (86.89) | 539 (224.31) |
| **2022/2023** | 1885 (678.58) | 81 (12.69) | 61 (5.94) | 328 (12.94) | 669 (87.44) | 539 (218.79) |
| **2023/2024†** | 1476 (531.35) | 71 (11.12) | 35 (3.41) | 170 (6.71) | 311 (40.65) | 305 (123.8) |

* Number of diagnosis is based on ICD-10 codes only, without laboratory confirmation
† For season 2023 - 2024 data was only available until 30 April 2024

**Supplemental table 3: Length of stay and hazard for discharge per age group for patients admitted with COVID-19, influenza and respiratory syncytial virus (RSV), Norway. 7 June 2022 – 30 April 2024. Including outliers.**

| | | | Length of Stay | | | | | | |
|---|---|---|---|---|---|---|---|---|---|
| Age group (Years of age) | Disease | Number of admitted patients* | Mean | Median | Q1 | Q3 | HR | 95 % Ci | p |
| Total | Covid-19 | 27116 | 6.89 | 4 | 2 | 7 | Ref. | - | - |
| | Influenza | 9552 | 4.85 | 3 | 1 | 5 | 1.35[*] | (1.31-1.38)[*] | <0.001[*] |
| | RSV | 5452 | 4.83 | 3 | 1 | 6 | 1.13[*] | (1.08-1.17)[*] | <0.001[*] |
| | Covid-19/Influenza | 507 | 5.89 | 3 | 2 | 6 | 1.10[*] | (1.00-1.20)[*] | 0.048[*] |
| | Covid-19/RSV | 233 | 7.25 | 4 | 2 | 7 | 0.88[*] | (0.77-1.00)[*] | 0.057[*] |
| | Influenza/RSV | 161 | 5.71 | 3 | 1 | 7 | 0.90[*] | (0.77-1.06)[*] | 0.220[*] |
| 0-4 | Covid-19 | 857 | 4.17 | 1 | 1 | 2 | Ref. | - | - |
| | Influenza | 739 | 2.87 | 1 | 1 | 3 | 1.48[†] | (1.31-1.68)[†] | <0.001[†] |
| | RSV | 3124 | 3.89 | 2 | 1 | 5 | 1.17[†] | (1.06-1.30)[†] | 0.003[†] |
| | Covid-19/Influenza | 19 | 2.47 | 2 | 1 | 4 | 1.47[†] | (0.88-2.44)[†] | 0.138[†] |
| | Covid-19/RSV | 90 | 4.47 | 2.5 | 1 | 5 | 1.02[†] | (0.81-1.30)[†] | 0.837[†] |
| | Influenza/RSV | 95 | 4.68 | 3 | 1 | 6 | 0.96[†] | (0.76-1.21)[†] | 0.701[†] |
| 5-14 | Covid-19 | 151 | 5.87 | 2 | 1 | 4 | Ref. | - | - |
| | Influenza | 579 | 2.93 | 1 | 1 | 3 | 2.00[†] | (1.55-2.57)[†] | <0.001[†] |
| | RSV | 137 | 3.74 | 2 | 1 | 4 | 1.75[†] | (1.28-2.40)[†] | <0.001[†] |
| | Covid-19/Influenza | 15 | 5.67 | 2 | 1 | 3 | 1.40[†] | (0.76-2.56)[†] | 0.281[†] |
| | Covid-19/RSV | 1 | 1.00 | 1 | 1 | 1 | 2.71[†] | (0.27-26.83)[†] | 0.394[†] |
| | Influenza/RSV | 9 | 7.33 | 4 | 1 | 9 | 0.62[†] | (0.27-1.42)[†] | 0.260[†] |
| 15-29 | Covid-19 | 658 | 4.59 | 2 | 1 | 4 | Ref. | - | - |
| | Influenza | 644 | 3.62 | 2 | 1 | 3 | 1.22[†] | (1.05-1.43)[†] | 0.011[†] |
| | RSV | 85 | 4.58 | 3 | 2 | 5 | 1.03[†] | (0.78-1.34)[†] | 0.858[†] |
| | Covid-19/Influenza | 32 | 5.38 | 1.5 | 1 | 3.5 | 0.86[†] | (0.58-1.28)[†] | 0.456[†] |
| | Covid-19/RSV | 5 | 4.40 | 2 | 1 | 2 | 1.22[†] | (0.49-3.06)[†] | 0.668[†] |
| | Influenza/RSV | 6 | 3.17 | 2 | 1 | 7 | 1.30[†] | (0.50-3.35)[†] | 0.593[†] |
| 30-64 | Covid-19 | 4967 | 7.91 | 3 | 1 | 7 | Ref. | - | - |
| | Influenza | 2685 | 4.64 | 2 | 1 | 5 | 1.69[†] | (1.59-1.79)[†] | <0.001[†] |
| | RSV | 455 | 6.47 | 4 | 2 | 6 | 1.26[†] | (1.14-1.40)[†] | <0.001[†] |
| | Covid-19/Influenza | 125 | 6.55 | 3 | 2 | 6 | 1.18[†] | (0.99-1.42)[†] | 0.068[†] |
| | Covid-19/RSV | 17 | 6.24 | 4 | 2 | 7 | 1.37[†] | (0.85-2.21)[†] | 0.201[†] |
| | Influenza/RSV | 18 | 6.39 | 3 | 1 | 4 | 1.25[†] | (0.78-2.02)[†] | 0.358[†] |
| 65-79 | Covid-19 | 10425 | 7.53 | 4 | 2 | 8 | Ref. | - | - |
| | Influenza | 2946 | 5.75 | 3 | 2 | 6 | 1.31[†] | (1.25-1.38)[†] | <0.001[†] |
| | RSV | 883 | 6.26 | 4 | 2 | 7 | 1.21[†] | (1.12-1.30)[†] | <0.001[†] |
| | Covid-19/Influenza | 187 | 5.73 | 3 | 2 | 7 | 1.21[†] | (1.04-1.41)[†] | 0.012[†] |
| | Covid-19/RSV | 67 | 11.12 | 5 | 3 | 13 | 0.73[†] | (0.57-0.93)[†] | 0.012[†] |
| | Influenza/RSV | 17 | 7.29 | 6 | 2 | 10 | 1.04[†] | (0.65-1.68)[†] | 0.866[†] |
| ≥80 | Covid-19 | 10058 | 6.13 | 4 | 2 | 7 | Ref. | - | - |
| | Influenza | 1959 | 5.50 | 4 | 2 | 7 | 1.10[†] | (1.04-1.16)[†] | 0.001[†] |
| | RSV | 768 | 6.28 | 4 | 3 | 7 | 0.95[†] | (0.87-1.03)[†] | 0.213[†] |
| | Covid-19/Influenza | 129 | 6.14 | 4 | 2 | 7 | 0.95[†] | (0.80-1.14)[†] | 0.611[†] |
| | Covid-19/RSV | 53 | 7.79 | 4 | 3 | 7 | 0.78[†] | (0.59-1.03)[†] | 0.078[†] |
| | Influenza/RSV | 16 | 9.44 | 6 | 2.5 | 13 | 0.69[†] | (0.42-1.13)[†] | 0.136[†] |

* Adjusted for the sex, age group of the admitted patient and Health region
† Adjusted for the sex of the admitted patient and Health region

**Supplemental table 4: Comparison of use of intensive care and/or ventilatory support (invasive mechanical ventilation, continuous positive airway pressure [CPAP] or bi-level positive airway pressure [BiPAP]) during the hospital stay, and number of deaths that occurred during admission or within 2 weeks of discharge among patients admitted with COVID-19, influenza and respiratory syncytial virus (RSV), Norway, 27 June 2022 – 30 April 2024. Including outliers**

| | | | Intensive care and respiratory support | | | | Deaths | | | |
|---|---|---|---|---|---|---|---|---|---|---|
| **Age group (Years of age)** | **Disease** | **Number of admitted patients*** | **n** | **OR** | **95% CI** | **p** | **n** | **OR** | **95% CI** | **p** |
| **TOTAL** | **Covid-19** | 27116 | 2196 (8.1) | Ref. | - | - | 2055 (7.6) | Ref. | - | - |
| | **Influenza** | 9552 | 755 (7.9) | 0.98 | (0.90-1.06) | 0.586 | 367 (3.8) | 0.49 | (0.44-0.55) | <0.001 |
| | **RSV** | 5452 | 983 (18.0) | 2.50 | (2.30-2.71) | <0.001 | 145 (2.7) | 0.34 | (0.28-0.40) | <0.001 |
| | **Covid-19/Influenza** | 507 | 54 (10.7) | 1.36 | (1.03-1.81) | 0.032 | 29 (5.7) | 0.75 | (0.52-1.09) | 0.133 |
| | **Covid-19/RSV** | 233 | 41 (17.6) | 2.43 | (1.73-3.41) | <0.001 | 14 (6.0) | 0.81 | (0.48-1.38) | 0.444 |
| | **Influenza/RSV** | 161 | 21 (13.0) | 1.74 | (1.10-2.75) | 0.017 | 5 (3.1) | 0.43 | (0.18-1.01) | 0.054 |
| **0-4** | **Covid-19** | 857 | 46 (5.4) | Ref. | - | - | 4 (0.5) | Ref. | - | - |
| | **Influenza** | 739 | 43 (5.8) | 1.08 | (0.70-1.65) | 0.731 | 2 (0.3) | 0.64 | (0.13-3.02) | 0.569 |
| | **RSV** | 3124 | 663 (21.2) | 4.69 | (3.44-6.38) | <0.001 | 2 (0.1) | 0.15 | (0.03-0.71) | 0.017 |
| | **Covid-19/Influenza** | 19 | 2 (10.5) | 2.48 | (0.63-9.73) | 0.192 | 0 (0.0) | - | - | - |
| | **Covid-19/RSV** | 90 | 17 (18.9) | 4.01 | (2.20-7.32) | <0.001 | 0 (0.0) | - | - | - |
| | **Influenza/RSV** | 95 | 16 (16.8) | 3.65 | (1.98-6.71) | <0.001 | 0 (0.0) | - | - | - |
| **5-14** | **Covid-19** | 151 | 22 (14.6) | Ref. | - | - | 2 (1.3) | Ref. | - | - |
| | **Influenza** | 579 | 40 (6.9) | 0.43 | (0.25-0.74) | 0.002 | 1 (0.2) | 0.15 | (0.02-1.18) | 0.072 |
| | **RSV** | 137 | 27 (19.7) | 1.44 | (0.78-2.66) | 0.246 | 3 (2.2) | 1.47 | (0.29-7.55) | 0.641 |
| | **Covid-19/Influenza** | 15 | 2 (13.3) | 0.95 | (0.23-3.97) | 0.948 | 0 (0.0) | - | - | - |
| | **Covid-19/RSV** | 1 | 0 (0.0) | - | - | - | 0 (0.0) | - | - | - |
| | **Influenza/RSV** | 9 | 1 (11.1) | 1.09 | (0.18-6.70) | 0.929 | 0 (0.0) | - | - | - |
| **15-29** | **Covid-19** | 658 | 33 (5.0) | Ref. | - | - | 5 (0.8) | Ref. | - | - |
| | **Influenza** | 644 | 38 (5.9) | 1.16 | (0.72-1.88) | 0.532 | 5 (0.8) | 0.97 | (0.30-3.18) | 0.958 |
| | **RSV** | 85 | 9 (10.6) | 2.18 | (1.02-4.68) | 0.045 | 1 (1.2) | 1.98 | (0.32-12.25) | 0.465 |
| | **Covid-19/Influenza** | 32 | 4 (12.5) | 2.79 | (0.97-8.05) | 0.058 | 0 (0.0) | - | - | - |
| | **Covid-19/RSV** | 5 | 1 (20.0) | 6.44 | (0.96-43.40) | 0.056 | 0 (0.0) | - | - | - |
| | **Influenza/RSV** | 6 | 0 (0.0) | - | - | - | 0 (0.0) | - | - | - |
| **30-64** | **Covid-19** | 4967 | 450 (9.1) | Ref. | - | - | 167 (3.4) | Ref. | - | - |
| | **Influenza** | 2685 | 213 (7.9) | 0.87 | (0.73-1.03) | 0.107 | 38 (1.4) | 0.41 | (0.29-0.59) | <0.001 |
| | **RSV** | 455 | 52 (11.4) | 1.31 | (0.96-1.77) | 0.086 | 13 (2.9) | 0.85 | (0.48-1.49) | 0.564 |
| | **Covid-19/Influenza** | 125 | 14 (11.2) | 1.30 | (0.75-2.28) | 0.350 | 3 (2.4) | 0.81 | (0.28-2.37) | 0.698 |
| | **Covid-19/RSV** | 17 | 1 (5.9) | 0.95 | (0.18-5.11) | 0.956 | 0 (0.0) | - | - | - |
| | **Influenza/RSV** | 18 | 2 (11.1) | 1.45 | (0.38-5.53) | 0.584 | 1 (5.6) | 2.41 | (0.45-12.98) | 0.307 |
| **65-79** | **Covid-19** | 10425 | 1083 (10.4) | Ref. | - | - | 748 (7.2) | Ref. | - | - |
| | **Influenza** | 2946 | 310 (10.5) | 1.01 | (0.88-1.15) | 0.908 | 132 (4.5) | 0.61 | (0.50-0.74) | <0.001 |
| | **RSV** | 883 | 153 (17.3) | 1.78 | (1.48-2.14) | <0.001 | 61 (6.9) | 0.96 | (0.73-1.26) | 0.757 |
| | **Covid-19/Influenza** | 187 | 21 (11.2) | 1.11 | (0.71-1.75) | 0.641 | 9 (4.8) | 0.68 | (0.35-1.32) | 0.259 |
| | **Covid-19/RSV** | 67 | 19 (28.4) | 3.39 | (2.00-5.77) | <0.001 | 7 (10.4) | 1.59 | (0.74-3.42) | 0.232 |
| | **Influenza/RSV** | 17 | 0 (0.0) | 1.00 | - | . | 0 (0.0) | 1.00 | - | . |
| **≥80** | **Covid-19** | 10058 | 562 (5.6) | Ref. | - | - | 1129 (11.2) | Ref. | - | - |
| | **Influenza** | 1959 | 111 (5.7) | 1.02 | (0.83-1.26) | 0.867 | 189 (9.6) | 0.85 | (0.72-1.00) | 0.051 |
| | **RSV** | 768 | 79 (10.3) | 1.96 | (1.53-2.52) | <0.001 | 65 (8.5) | 0.77 | (0.59-0.99) | 0.045 |
| | **Covid-19/Influenza** | 129 | 11 (8.5) | 1.63 | (0.88-3.00) | 0.119 | 17 (13.2) | 1.24 | (0.75-2.07) | 0.400 |
| | **Covid-19/RSV** | 53 | 3 (5.7) | 1.18 | (0.40-3.50) | 0.765 | 7 (13.2) | 1.30 | (0.60-2.82) | 0.514 |
| | **Influenza/RSV** | 16 | 2 (12.5) | 2.96 | (0.77-11.38) | 0.114 | 4 (25.0) | 2.97 | (1.00-8.83) | 0.050 |

**Supplemental table 5: Pearson correlation of case definitions using ICD-10 codes for severe acute respiratory infections (SARI) in combination with laboratory confirmation compared to disease specific ICD-10 codes, Norway, 26 June 2020 – 30 April 2024**

| Age group | Pearson correlation of RSV case definitions (P-value) | Additional cases identified using SARI + laboratory confirmation of RSV, compared to RSV specific ICD-10 codes | Pearson correlation of Influenza case definitions (P-value) | Additional cases identified using SARI + laboratory confirmation of Influenza, compared to Influenza specific ICD-10 codes |
|---|---|---|---|---|
| **0-4** | 0.999 (<0.001) | 253 | 0.992 (<0.001) | -24 |
| **5-14** | 0.954 (<0.001) | 89 | 0.992 (<0.001) | 19 |
| **15-29** | 0.956 (<0.001) | 55 | 0.994 (<0.001) | -21 |
| **30-64** | 0.991 (<0.001) | 161 | 0.999 (<0.001) | -52 |
| **65-79** | 0.991 (<0.001) | 218 | 0.998 (<0.001) | 12 |
| **≥80** | 0.990 (<0.001) | 124 | 0.999 (<0.001) | -33 |
| **Total** | 0.999 (<0.001) | 900 | 0.999 (<0.001) | -123 |
